# Testing the causal relationships of physical activity and sedentary behaviour with mental health and substance use disorders: A Mendelian Randomisation study

**DOI:** 10.1101/2022.07.31.22278200

**Authors:** Eleonora Iob, Jean-Baptiste Pingault, Marcus R. Munafò, Brendon Stubbs, Mark Gilthorpe, Adam X. Maihofer, Psychiatric Genomics Consortium Posttraumatic Stress Disorder Working Group, Andrea Danese

## Abstract

**Importance:** Observational studies suggest that physical activity can reduce the risk of mental health and substance use disorders. However, it is unclear whether this relationship is causal or explained by confounding (e.g., common underlying causes or reverse causality).

**Objective:** We investigated bidirectional causal relationships of physical activity (PA) and sedentary behaviour (SB) with mental health and substance use disorders, applying a genetically informed causal inference method.

**Design, Setting, and Participants:** This two-sample Mendelian Randomisation (MR) study used genetic instruments for the exposures and outcomes that were derived from the largest available, non-overlapping genome-wide association studies (GWAS). Summary-level data for objectively assessed PA (accelerometer-based average activity, moderate activity, and walking) and SB (assessed over 7 consecutive days) and self-reported moderate-to-vigorous PA were obtained from the UK Biobank. Data for mental health/substance use disorders were obtained from the Psychiatric Genomics Consortium and the GWAS and Sequencing Consortium of Alcohol and Nicotine Use. MR estimates were combined using inverse variance weighted meta-analysis (IVW). Several sensitivity analyses were conducted to assess the robustness of the results (e.g., MR-Egger, weighted median/mode, MR-RAPS, MR-PRESSO).

**Exposures:** Objectively assessed/self-reported PA and objectively assessed SB.

**Main Outcomes and Measures:** Mental health and substance use disorders.

**Results:** Accelerometer-based average PA had a causal protective effect on the risk of depression (b=-0.043, 95%CI: -0.071 to -0.016, effect size[OR]=0.957), and on the number of cigarettes smoked per day (b=-0.026; 95%CI: -0.035 to -0.017, effect size[β]=-0.022). Accelerometer-based SB was causally related to a lower risk of anorexia (b=-0.341, 95%CI: -0.530 to -0.152, effect size[OR]=0.711) and schizophrenia (b=-0.230; 95%CI: -0.285 to -0.175, effect size[OR]=0.795). However, we found evidence of reverse causality in the effect of SB on schizophrenia. Further, PTSD, bipolar disorder, anorexia, and ADHD were all causally related to increased PA.

**Conclusions and Relevance:** This study provides evidence consistent with a causal protective effect of objectively assessed but not self-reported PA on reduced depression and cigarette smoking. Objectively assessed SB had a protective effect on anorexia. Enhancing PA may be an effective prevention strategy for specific types of psychiatric disorders.

**KEY POINTS:** *Question:* Do heightened physical activity and low sedentary behaviour have causal protective effects on the risk of mental health and substance use disorders?

*Findings:* Applying two-sample Mendelian Randomisation to summary-level data from large-scale genome-wide association studies to strengthen causal inferences, we found evidence that objectively assessed but not self-reported physical activity was causally related to a lower risk of depression and cigarette smoking, whereas sedentary behaviour had a protective effect on anorexia.

*Meaning:* Interventions that enhance physical activity may be effective in reducing the risk of depression and cigarette smoking, whereas those that increase sedentary behaviours may be effective to reduce the risk of anorexia nervosa.

## INTRODUCTION

Mental health and substance use disorders affect around one in three people across the lifespan^1^ and are leading causes of the global burden of disease and disability.^2,3^ Rates of common mental disorders, such as depression and anxiety, are increasing among children and young people,^4^ indicating little improvement in the efficacy or implementation of current preventive strategies. Furthermore, despite several advances in psychological and pharmacological interventions, many individuals do not respond well to standard treatments,^5^ which also do not address the recognised physical burden of mental illness.^6^ Hence, novel approaches are necessary in order to prevent and treat psychiatric disorders.^7^

A growing body of evidence suggests that enhancing physical activity levels may be an effective strategy to prevent and treat mental health and substance use disorders.^7,8^ Meta-analyses examining the prospective relationships of physical activity with mental health in the population have found that higher levels of physical activity may offer protection against the onset of depression,^9^ stress-related disorders,^10,11^ and psychotic disorders.^12^ Correspondingly, prospective studies have also shown that high levels of sedentary behaviour are associated with an increased risk of these disorders.^13–15^ Furthermore, meta-analyses of randomised controlled trials (RCTs) have provided evidence of the efficacy of physical activity interventions to reduce mental health symptoms and improve neurocognitive outcomes among individuals affected by depression, stress-related disorders, and schizophrenia.^16^ Beyond mental health outcomes, research has also highlighted the potential beneficial role of physical activity in preventing and reducing substance use problems.^17^ Observational studies suggest that physical inactivity and sedentary behaviour are linked to an increased risk of alcohol consumption and cigarette smoking.^18–20^ Additionally, meta-analyses of clinical studies have found that physical exercise can effectively increase abstinence rates, reduce craving and withdrawal symptoms, and ameliorate psychological wellbeing and quality of life in people with substance use disorders.^21,22^

Despite this evidence, it is unclear whether physical activity is causally related to the risk of mental health and substance use disorders, or whether this relationship might be better explained by reverse causation and/or common causes. Although RCTs are considered the gold standard approach for establishing causality, these studies have predominantly tested the remedial effects of physical activity in at-risk samples, rather than testing its protective effects in the general population. RCTs are also limited by strict inclusion criteria and close monitoring of the participants’ adherence to the treatment protocol, and therefore their results might not be generalisable to real-world settings. In contrast, observational prospective studies are ideally suited for studying the real-world protective effects of physical activity on psychiatric disorders. However, due to the lack of randomisation, a variety of social, behavioural, and genetic factors could be associated with both physical activity and mental health, thereby potentially acting as confounders of their relationship. Observational studies to date have only considered a limited set of confounders, and the associations between physical activity and mental health are usually weaker when confounding factors are taken into account. Furthermore, physical activity could have bidirectional links with mental health and substance use disorders. For instance, some research suggests that mental disorders such as depression can lead to the development of sedentary behaviour and decreased levels of physical activity.^23^ However, only few studies have investigated both temporal relationships simultaneously. There is also limited evidence from well-designed prospective studies or RCTs on the relationship of physical activity with bipolar disorder and developmental disorders such as attention deficit hyperactivity disorder (ADHD), autism, and eating disorders. Lastly, it is unclear whether the measurement (i.e., self-reported vs objectively assessed) and intensity of physical activity may also play a role. Research to date has predominantly used self-reported measures of physical activity, which might not accurately capture specific levels of intensity and are particularly prone to confounding by cognitive function, mood, and social desirability biases.^24^

Mendelian randomisation (MR) is a genetically informed method that uses genetic variants associated with an exposure as instrumental variables for investigating causal relationships with the outcome and vice versa.^25^ This approach can reduce confounding effects since genetic variants are thought to be randomly distributed at conception, do not change over time, and cannot be affected by disease status. Using this approach, two studies have found that objective accelerometer-based physical activity had a protective causal relationship with depression and bipolar disorder.^26,27^ However, no study to date has used this approach to test the bidirectional relationships of physical activity and sedentary behaviour with multiple diagnoses of mental health and substance use disorders.

The present study aimed to (i) investigate the causal nature of the relationship of physical activity and sedentary behaviour with mental health and substance use disorders, and (ii) shed light on the direction of this relationship. We applied two-sample MR in order to test bidirectional associations of physical activity and sedentary behaviour with mental health and substance use disorders based on results from large genome-wide association studies (GWASs), using both self-reported and objective accelerometer-based physical activity data.

## METHODS

### Study design

A two-sample MR design was used to test bidirectional pathways between physical activity and mental health and substance use disorders. The analyses were conducted with physical activity as (i) the exposure, to assess whether it has a causal effect on mental health/substance use disorders, and as (ii) the outcome, to assess whether mental health/substance use disorders have a causal effect on physical activity. Summary-level data for all exposure and outcome variables were derived from large-scale, non-overlapping GWASs in individuals of European ancestry. We considered five different physical activity exposures in order to evaluate the role of different assessment methods and intensity levels. These included self-reported moderate-to-vigorous activity and accelerometer-based average activity (i.e., mean acceleration), moderate activity, walking, and sedentary behaviour. An outcome-wide approach was adopted in order to assess the causal effects of physical activity on several individual disorders, including depression, post-traumatic stress disorder (PTSD), bipolar disorder, schizophrenia, anorexia nervosa, ADHD, autism, alcohol dependence, cannabis use disorder, and cigarette smoking. We also included birth length as a negative control outcome, as it is impossible that physical activity levels affect perinatal outcomes. Figure1 provides an overview of the study design and the core MR assumptions for valid instrumental variables.

**Figure1.**
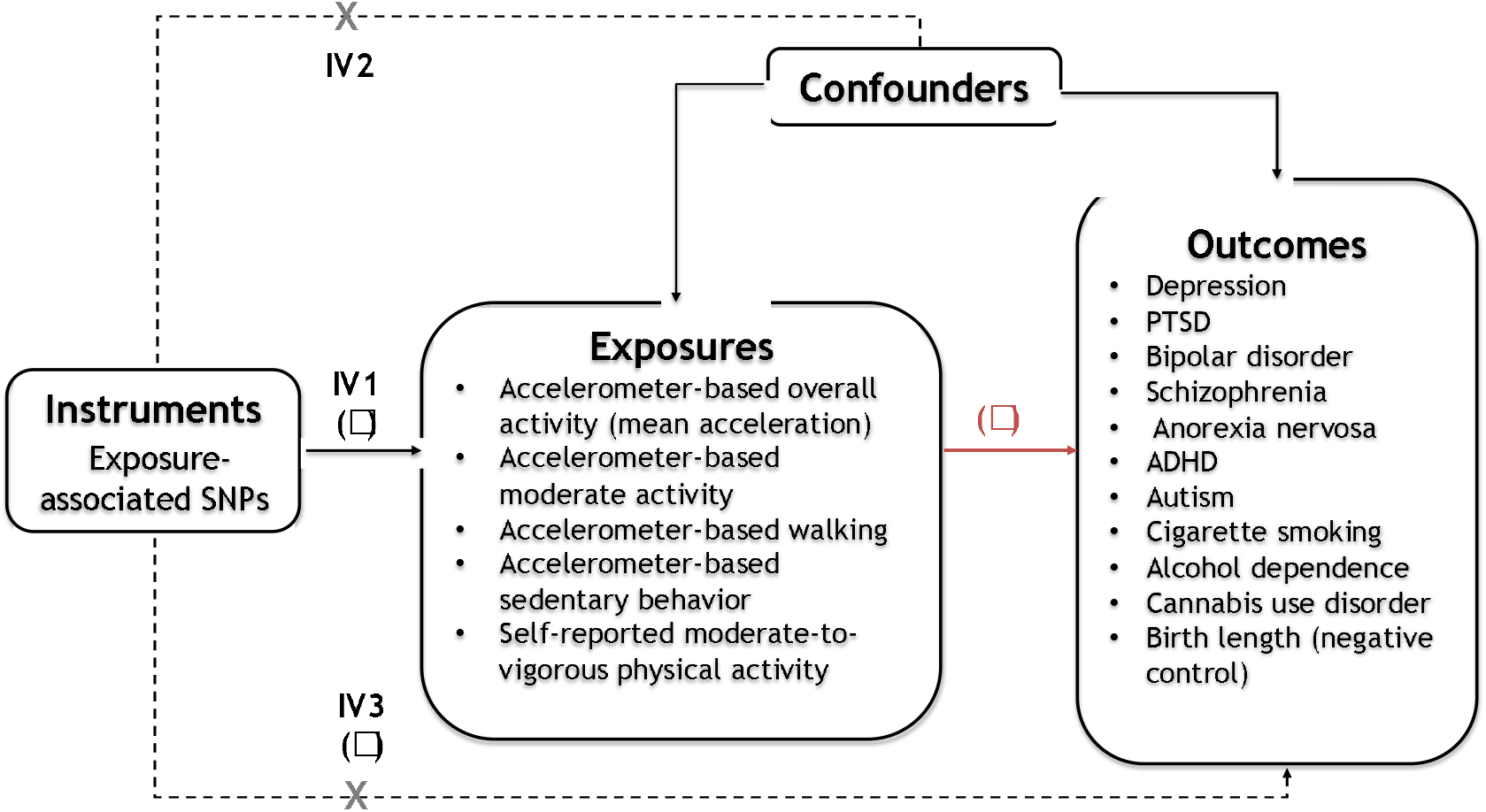
Study design and Mendelian Randomisation (MR) assumptions. **Note**. (a) Study design: Solid paths are hypothesised to exist, whereas dotted paths are hypothesised not to exist according to MR assumptions; *β* is the causal relationship of interest to be estimated, where *β* = *γ*/*α*. *γ* and *α* are the estimated direct effects of a SNP on the exposure and outcome. (b) MR assumptions: MR relies on three core assumptions for valid instrumental variables. These include: Relevance (IV1) – the instrument is associated with the risk factor of interest; Exchangeability (IV2) – the instrument is not associated with any potentially confounding variable; and Exclusion Restriction (IV3) – the instrumental variable can only influence the outcome via the risk factor (Figure 1). In light of the first assumption, the genetic instruments were constructed using top SNPs associated with the exposure variables. The second and third assumptions are violated if instrument SNPs show horizontal pleiotropy, influencing the outcome through other causal pathways than the exposure, or correlated pleiotropy, where genetic variants for the exposure are also associated with a confounder.^25^ Therefore, several sensitivity analyses were conducted to detect and remove possible pleiotropic genetic variants, as detailed in the Methods and Results. SNP = single nucleotide polymorphism.

### GWAS data sources

#### (i) Physical activity and sedentary behaviour

Summary statistics for self-reported moderate-to-vigorous activity (N∼377,000) and accelerometer-based average activity, moderate activity, walking, and sedentary behaviour (N∼91,000) were obtained from the UK Biobank.^28,29^ Self-reported moderate-to-vigorous activity during work and leisure time was calculated as the sum of total minutes per week of moderate activity (e.g., carrying light loads, cycling at normal pace) multiplied by four and the total minutes per week of vigorous activity (e.g., fast cycling, aerobics, heavy lifting) multiplied by eight in order to reflect their metabolic equivalents.^28^ To objectively assess physical activity, UK Biobank participants were invited to wear a wrist-worn accelerometer at all times for 7 days. Levels of activity were measured in milli-gravity units (mg). The accelerometer data were then used to derive different phenotypes representing average activity, moderate activity, walking, and sedentary activity, which were defined using machine learning algorithms.^29^

#### (ii) Mental health and substance use disorders

Summary statistics for diagnoses of major depressive disorder^30^ (N∼143,000), PTSD (N∼956,000) (Freeze 3, Nievergelt et al., in prep.), bipolar disorder^31^ (N∼413,000), schizophrenia^32^ (N∼306,000), anorexia nervosa^33^ (N∼69,000), ADHD^34^ (N∼55,000), autism^35^ (N∼46,000), alcohol dependence^36^ (N∼47,000), and cannabis use disorder^37^ (N∼374,000) were obtained from the Psychiatric Genomics Consortium (PGC). Summary statistics for cigarette smoking^38^ (i.e., number of cigarettes smoked per day) (N∼143,000) were obtained from the GWAS and Sequencing Consortium of Alcohol and Nicotine Use (GSCAN), and those for birth length^39^ (i.e., sex- and age-adjusted standardised scores) (N∼28,000) from the Early Growth Consortium (EGC). We used meta-analytic results that left out UK Biobank participants for depression, PTSD, bipolar disorder, anorexia nervosa, and cigarette smoking in order to avoid sample overlap between the exposure and outcome data. For depression, we also excluded participants from 23andMe owing to access constraints.

Further information regarding the data sources, sample size, and instrument strength of the included GWAS datasets can be found in Appendix 1 (eMethods, eTable1). All original studies included in the GWAS datasets have been granted ethical approval.

### Selection of genetic instruments

We created two sets of genetic instruments for each exposure variable; the first set (G1) included only SNPs reported as genome-wide significant (p<5×10^−8^), and the second set (G2) included top SNPs meeting a more relaxed threshold (p<1×10^−6^). This approach of relaxing the genome-wide significance threshold for genetic instruments has been previously used in psychiatric MR research.^27^ SNPs that were correlated at r^2^>0.001 were clumped to ensure independence between the genetic variants included as instruments. SNPs for the exposure that were not available in the summary statistics of the outcome were replaced with overlapping proxy SNPs in high-linkage disequilibrium (r^2^>0.8). The resulting list of SNPs used as instruments for each phenotype is shown in Appendix 2 (eTable2).

### Statistical analyses

We considered physical activity/sedentary behaviour and mental health/substance use disorders as exposures in turn to assess potential bidirectional pathways between these. As the primary analysis, we used random-effects inverse-variance weighted (IVW) regression to combine effect estimates (i.e., Wald ratios) from multiple SNPs. For genetic instruments involving a single SNP, individual Wald ratios (WR) are presented instead. As measures of effect size, odds ratios (OR) are reported for binary outcomes and standardised beta coefficients (β)^40^ for continuous outcomes. Given the large number of tests performed, we calculated false discovery rate (FDR) corrected p-values to account for the multiple exposures and outcomes used to test each direction of causation (55 tests in total). In sensitivity analyses, a variety of robust MR methods were used to identify and correct for potential violations of key MR assumptions, including MR-Egger, weighted median, weighted mode, MR-PRESSO, MR-RAPS, and Steiger directionality test and filtering. Additionally, we conducted Cochran’s (IVW) and Rucker’s (MR Egger) Q tests to detect heterogeneous causal effects when using meta-analytic methods. An overview of the MR methods and the rationale for their application in our study is provided in Table1. All statistical analyses were conducted in R (version 4.0.2) using the *TwoSampleMR* package.^41^ The study protocol was pre-registered in the Open Science Framework (OSF) (https://osf.io/ceptf), and any deviations that have occurred from our preregistered plans are outlined in Appendix 1 (eMethods).

**Table1.** Description of the MR methods used in the main and sensitivity analyses.

| MR method | General description | Rationale for application | Assumptions/limitations |
| --- | --- | --- | --- |
| *Main analyses* |  |  |  |
| Wald ratio | Ratio of the effect of the SNP-outcome association by the SNP-exposure association. | Main MR method used for genetic instruments including a single SNP. | Provides valid estimates if the genetic instrument satisfies all IV assumptions. |
| Inverse-variance weighted (IVW) regression<sup>49</sup> | Linear regression of the SNP-outcome associations on the SNP-exposure associations, weighted by the inverse-variance of the SNP-outcome associations and with intercept constrained to zero. | Main MR method used to combine effect estimates for genetic instruments including $\geq 2$ SNPs. | Provides valid estimates if the genetic instrument satisfies all IV assumptions.<br>Accounts for balanced pleiotropy (i.e., average pleiotropic effect equals to zero), but susceptible to unbalanced pleiotropy (i.e., average pleiotropic effect is positive or negative). |
| *Sensitivity analyses* |  |  |  |
| MR-Egger regression<sup>50</sup> | Weighted linear regression similar to IVW, but with intercept unconstrained. | Provides an estimate of unbalanced horizontal pleiotropy and can yield accurate MR estimates even if all instruments are invalid.<br>The intercept represents the average unbalanced horizontal pleiotropic effect across SNPs. | Makes IV1, IV2, and InSIDE assumptions (i.e., the SNP-exposure associations are independent of the direct effects of the genetic variants on the outcome).<br>Relaxes IV3 assumption.<br>But suffers from low power and is sensitive to outliers. |
| Weighted median method<sup>51</sup> | Weighted median estimator for combining effect estimates from multiple genetic variants (instead of weighted mean as in IVW). | The median of effect estimates is more robust to outliers than the corresponding mean (pleiotropy often manifests in the presence of genetic variants with outlying effect estimates).<br>Provides accurate MR estimates when the majority of the information (>50%) comes from valid instruments. | Makes IV1 and IV2 assumptions.<br>Relaxes IV3 assumption. |
| Weighted mode method<sup>52</sup> | Weighted mode estimator for combining effect estimates from multiple genetic variants (instead of weighted mean as in IVW). | Like the median, the mode is more robust to outliers than the corresponding mean.<br>Provides accurate MR estimates if the largest subset of SNPs with a similar effect ratio (i.e., mode) is formed by valid instruments, even if the<br>majority of SNPs are invalid. | Makes IV1 and IV2 assumptions.<br>Relaxes IV3 assumption. |
| MR-PRESSO<sup>53</sup> | Performs 3 tests: (1) detection of horizontal pleiotropy (global test); (2) correction for horizontal pleiotropy by removal of outliers (outlier test); (3) test for significant differences in the MR estimates before and after outlier removal (distortion test). | Identifies and removes horizontal pleiotropic outliers in instruments including multiple SNPs. | Makes IV1 and IV2 assumptions.<br>Relaxes IV3 assumption.<br>Best suited when horizontal pleiotropy occurs in < 50% of instruments. |
| Robust adjusted profile score (MR-RAPS)<sup>54</sup> | SNPs are assigned different weights according to the strength of their associations. | Allows for the use of weaker instruments, which is not recommended for other methods.<br>In our study, MR-RAPS is only used for the G2 instruments, which have been constructed using a more liberal p-value threshold and are therefore more susceptible to weak instrument bias. | Makes IV2 and IV3 assumptions.<br>Relaxes IV1 assumption. |
| Steiger directionality test and filtering<sup>55</sup> | Steiger Z-test assesses whether the absolute correlation of the genetic variants with the exposure is larger than that with the outcome.<br>If Z-value > 0, X causes Y; if Z-value < 0, Y causes X; if Z = 0, neither direction is accepted.<br>Steiger filtering can then be used to correct for potential misspecification of the direction of effect by removing genetic variants that explain more variation in the outcome than the exposure. | Indicates the direction of the causal association (sign of Z-value) and the confidence level of the direction (p-value).<br>Identifies and removes genetic variants whose direction of effect has been misspecified. | Results may be biased in the presence of horizontal pleiotropy or different levels of measurement error between the exposure and the outcome. |
| **Note.** MR = Mendelian randomisation; SNP = single nucleotide polymorphism; IV = instrumental variable. |  |  |  |

## RESULTS

The results of the main MR analyses (IVW/WR) are illustrated in Figures2-3. The sensitivity analyses with MR-Egger, weighted median, weighted mode, and MR-RAPS are shown in eFigures1-2 (Appendix 1) and are also reported in eTables3-6 (Appendix 2). The results of other sensitivity analyses, including MR-Egger intercept, Q statistics, MR-PRESSO, and Steiger directionality test/filtering, are shown in eTable2 and eTables7-11 (Appendix 2). In the following sections, we focus on the results that were robust to the correction for multiple testing (i.e., FDR-adjusted p-value < 0.05), which are also reported in Table2.

**Figure2.**
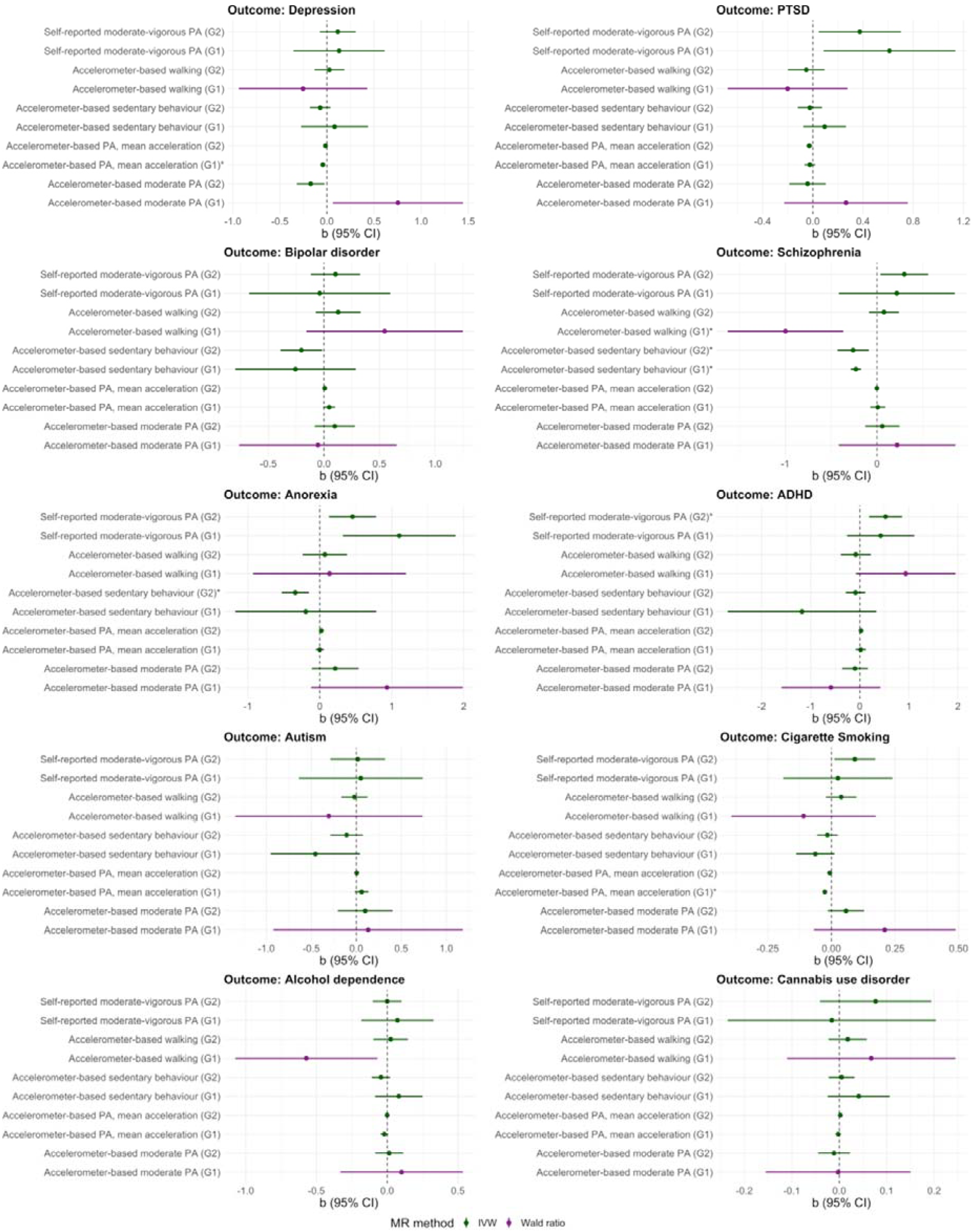
MR estimates (IVW/Wald ratio) and 95% confidence intervals for the causal relationships of physical activity and sedentary behaviour with mental health and substance use disorders (Direction 1). **Note**. MR = Mendelian randomisation; IVW = inverse variance weighted; G1 = genome-wide significant genetic instrument (P < 5×10^−8^); G2 = more relaxed genetic instrument (P < 1×10^−6^); PA = physical activity. IVW is used for analyses involving ≥ 2 SNPs, and Wald ratio for analyses involving 1 SNP. Effects marked with an asterisk (*) are robust to the correction for multiple testing (i.e., FDR-adjusted p-value < 0.05).

**Figure3.**
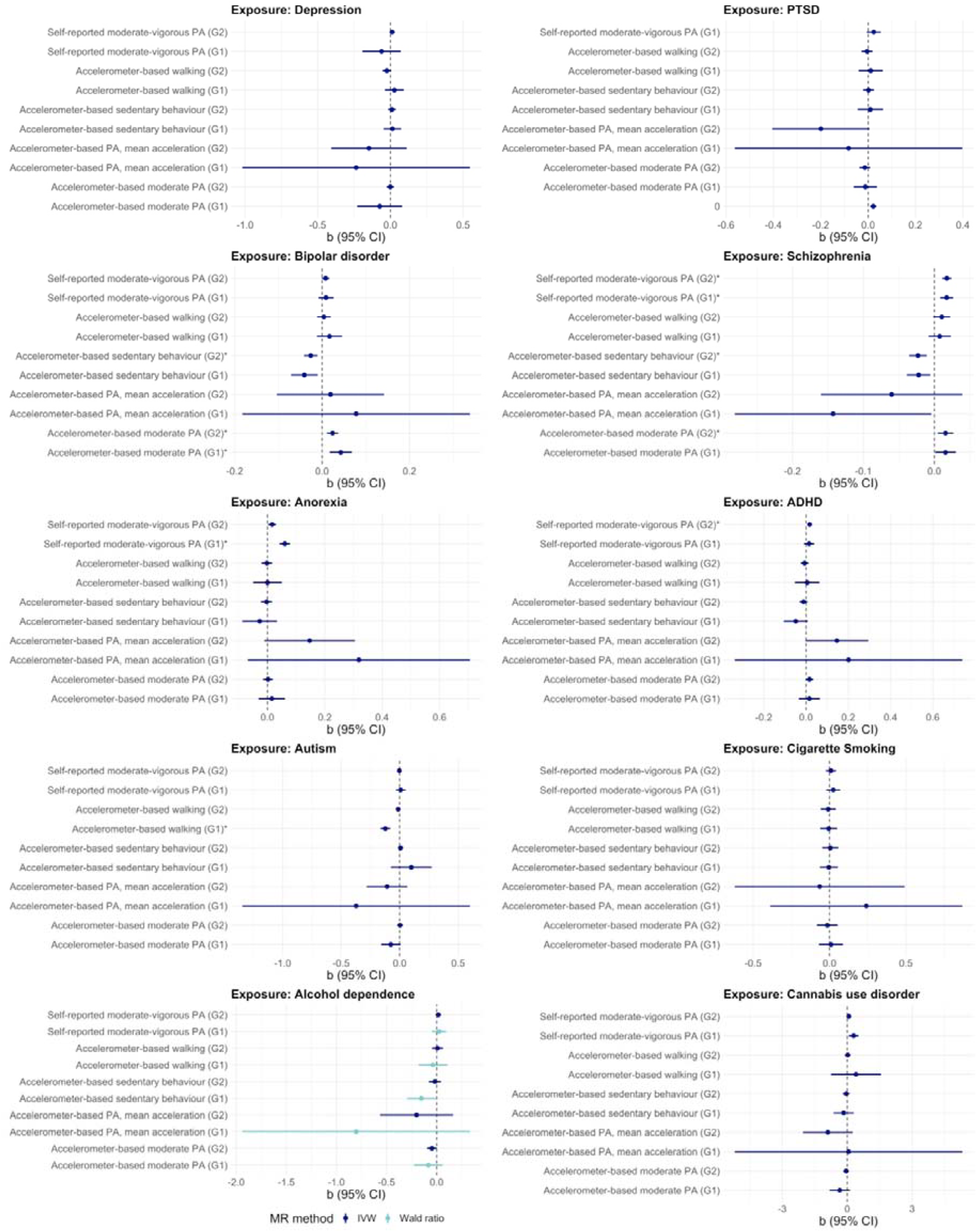
MR estimates (IVW/Wald ratio) and 95% confidence intervals for the causal relationships of mental health and substance use disorders with physical activity and sedentary behaviour (Direction 2). **Note**. MR = Mendelian randomisation; IVW = inverse variance weighted; G1 = genome-wide significant genetic instrument (P < 5×10^−8^); G2 = more relaxed genetic instrument (P < 1×10^−6^); PA = physical activity. IVW is used for analyses involving ≥ 2 SNPs, and Wald ratio for analyses involving 1 SNP. Effects marked with an asterisk (*) are robust to the correction for multiple testing (i.e., FDR-adjusted p-value < 0.05).

**Table2.**
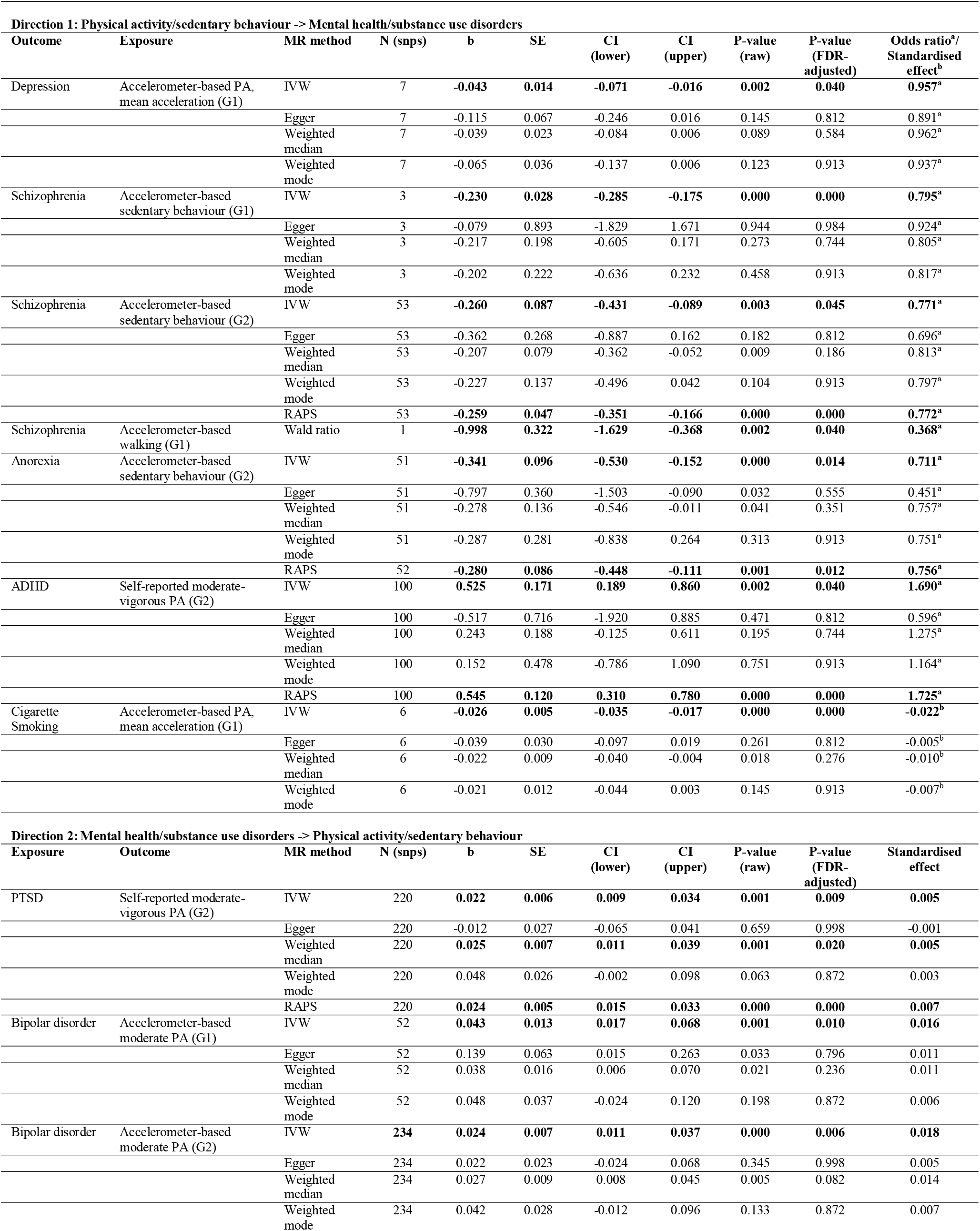
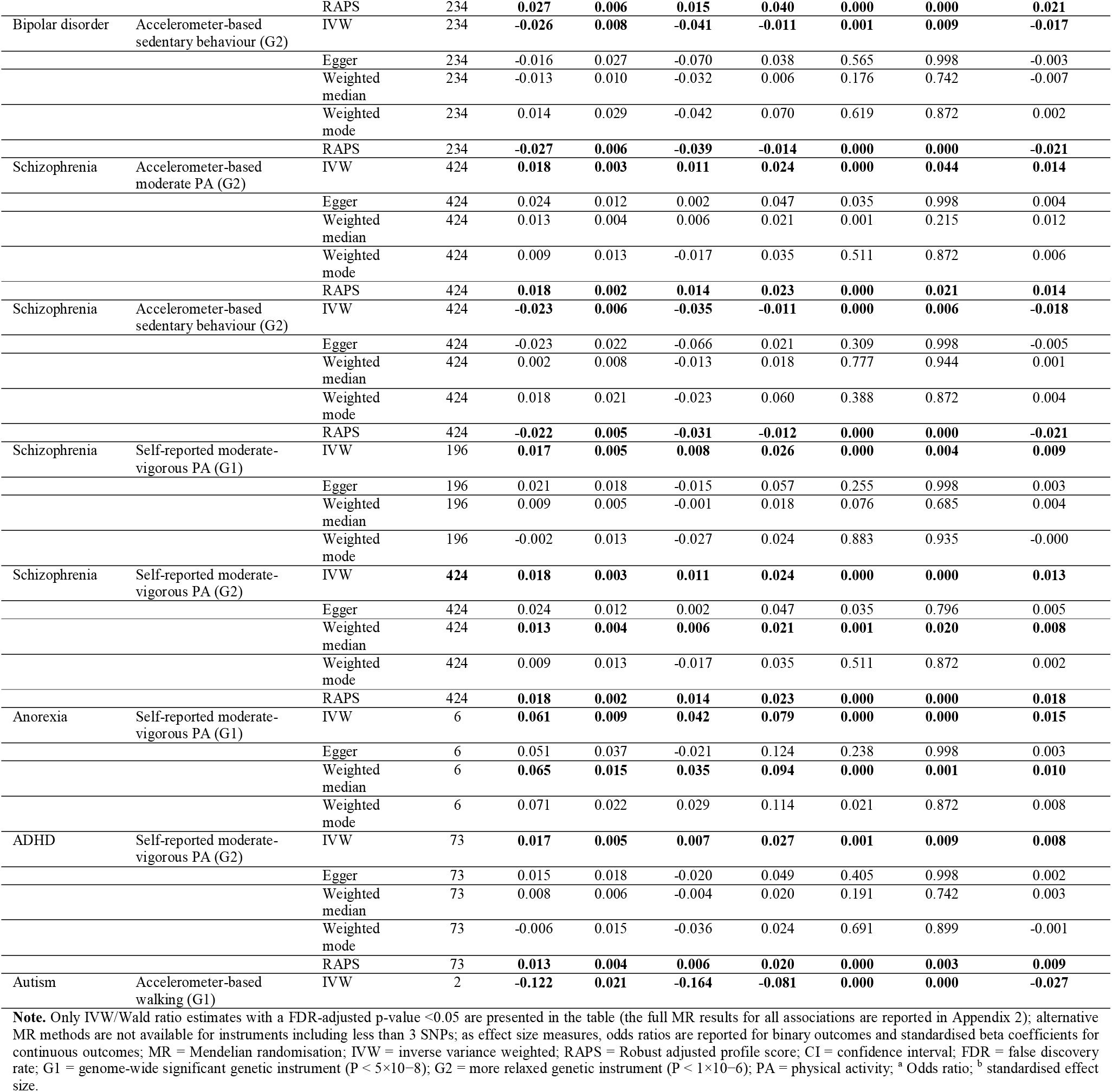
Main MR results and sensitivity analyses for the bidirectional relationships of physical activity and sedentary behaviour with mental health and substance use disorders.

### Main analyses

#### Direction 1: Effects of physical activity/sedentary behaviour on mental health/substance use disorders

Higher levels of genetically predicted accelerometer-based average physical activity had a protective effect on depression (G1: IVW b=-0.043, 95%CI: -0.071 to -0.016) and cigarette smoking (G1: IVW b=-0.026, 95%CI: -0.035 to -0.017). Genetically predicted accelerometer-based sedentary behaviour was causally related to a lower risk of schizophrenia at both instrument thresholds (G1: IVW b=-0.230, 95%CI: -0.285 to -0.175; G2: IVW b=-0.260, 95%CI: -0.431 to -0.089), and accelerometer-based walking had a protective effect on schizophrenia (G1: WR b=-0.998, 95%CI: -1.629 to -0.368). However, the latter effect was driven by a single SNP, and its direction was inconsistent when using the more relaxed instrument threshold. Sedentary behaviour also had a protective effect on anorexia nervosa (G2: IVW b=-0.341, 95%CI: -0.530 to -0.152). Higher levels of self-reported moderate-to-vigorous activity increased the risk of ADHD (G2: IVW b=0.525, 95%CI: 0.189 to 0.860) (Figure2). The odds ratios of these effects ranged from small to moderate^42^ (Table2). As expected, we found weak evidence of a causal effect of physical activity on birth length (i.e., negative control outcome; eTable3).

#### Direction 2: Effects of mental health/substance use disorders on physical activity/sedentary behaviour

Genetically predicted PTSD was causally related to higher levels of self-reported physical activity (G2: IVW b=0.022, 95%CI: 0.009 to 0.034). Genetically predicted bipolar disorder was causally related to lower levels of sedentary behaviour (G2: IVW b=-0.026, 95%CI: -0.041 to -0.011), and we also observed a positive causal effect of bipolar disorder on accelerometer-based moderate activity at both instrument thresholds (G1: IVW b=0.043, 95%CI: 0.017 to 0.068, p-value=0.001; G2: IVW b=0.024, 95%CI: 0.011 to 0.037, p<0.001). Genetically predicted schizophrenia was causally linked to lower levels of accelerometer-based sedentary behaviour (G2: IVW b=-0.023, 95%CI: -0.035 to -0.011), to higher levels of accelerometer-based moderate activity (G2: IVW b=0.018, 95%CI: 0.011 to 0.024), and to higher levels of self-reported moderate-to-vigorous physical activity (G1: IVW b=0.017, 95%CI: 0.008 to 0.026; G2: IVW b=0.018, 95% CI: 0.011 to 0.024). Genetically predicted anorexia (G1: IVW b=0.061, 95%CI: 0.042 to 0.079) and ADHD (G2: IVW b=0.017, 95%CI: 0.007 to 0.027) were causally related to higher levels of self-reported physical activity. Genetically predicted autism had a causal effect on reduced levels of accelerometer-based walking (G1: IVW b=-0.122, 95%CI: -0.164 to -0.081) (Figure3). However, the size of these effects was generally small (Table2).

### Sensitivity analyses

The results of the sensitivity analyses with MR-Egger, weighted median, and weighted mode revealed effects in the same direction as those observed in the main analyses, but the confidence intervals were often imprecise (Table2). Of note, these sensitivity methods have lower statistical power than IVW because they rely on stricter assumptions, and therefore their results are expected to provide weaker statistical evidence but not effect sizes. MR-RAPS provided consistent and precise results across most outcomes (Table2). The intercept of MR-Egger (eTable8), Q statistics (eTable7), and MR-PRESSO (eTable10) provided little evidence of heterogeneity and unbalanced horizontal pleiotropy in the instrument effects of physical activity/sedentary behaviour on depression, anorexia, and cigarette smoking and in the instrument effects of anorexia on physical activity. In contrast, Q statistics and MR-PRESSO tests highlighted the presence of heterogeneous causal estimates and outliers in the G2 instrument effects of sedentary behaviour and self-reported physical activity on depression and ADHD, respectively, and in the effects of PTSD, bipolar disorder, schizophrenia, and ADHD on physical activity/sedentary behaviour. These effects were generally smaller and more precise following the removal of outliers by MR-PRESSO. The Steiger test for the average effect of all the genetic variants associated with a particular phenotype suggested that the overall direction of the observed MR effects was correct. When considering the effects of individual SNPs, we found evidence of misspecified SNPs in the MR analysis of self-reported physical activity and ADHD, and the causal effect was considerably smaller and no longer consistent with one direction after their removal by Steiger filtering, thereby suggesting that self-reported physical activity was not causally related to ADHD. We also observed misspecified SNPs in the instruments for bipolar disorder and schizophrenia, but the magnitude and precision of their causal effects did not change substantially after applying Steiger filtering (eTable11).

## DISCUSSION

Using data from large-scale GWASs, we applied two-sample MR to test whether physical activity and sedentary behaviour are causally related to mental health and substance use disorders, or vice versa. The results showed that objectively assessed but not self-reported physical activity had a causal protective effect on the risk of depression and cigarette smoking. In contrast, objectively assessed sedentary behaviour had a protective effect on anorexia and schizophrenia, and objectively assessed walking reduced the risk of schizophrenia. We also found evidence suggesting a causal effect of mental health disorders on physical activity/ sedentary behaviour. PTSD, schizophrenia, anorexia, and ADHD were all causally related to higher levels of self-reported physical activity. Schizophrenia and bipolar disorder had a causal relationship with higher levels of objectively assessed moderate activity and with reduced levels of sedentary behaviour, whereas autism was linked to lower walking activity. These findings highlight the important but complex nature in which physical activity and sedentary behaviour are related to mental and substance use disorders.

### Protective effect of physical activity on depression and cigarette smoking

In line with the results of earlier observational studies^9^, we found evidence of a 5% reduction in the odds of depression for every 1 standard deviation (SD) increase in objectively assessed average activity. Our results also extend previous findings by showing that other types/intensity levels of physical activity (i.e., objectively assessed moderate activity and walking, self-reported physical activity) and sedentary behaviour were not causally related to depression, suggesting that increasing average levels of physical activity may be the most effective strategy to reduce the risk of depression. Earlier studies have provided mixed findings regarding the impact of physical activity interventions on smoking cessation^43^, and little is currently known regarding the preventive effects of physical activity on smoking initiation or the levels of smoking among smokers. Our results suggest that every 1 SD increase in objectively assessed average activity may result in 0.26 fewer cigarettes smoked per day. In the opposite direction of causation, we found weak evidence for the causal effects of depression and cigarette smoking on physical activity/sedentary behaviour, suggesting that the relationships of physical activity/sedentary behaviour with depression and cigarette smoking are unlikely to be explained by reverse causality.

### Causal pathways between physical activity and schizophrenia, PTSD, and bipolar disorder

Contrary to earlier evidence suggesting that physical activity can reduce the risk of psychotic disorders,^12^ our results indicate that a 1 SD increase in the amount of sedentary behaviour can reduce the odds of schizophrenia by 20%. We also observed a 60% reduction in the risk of schizophrenia for every 1 SD increase in walking activity. However, in the opposite direction of causation, schizophrenia was causally related to lower levels of sedentary behaviour, as well as being associated with higher levels of objectively assessed moderate activity and self-reported physical activity, and therefore reverse causality might be at play. These results outline the complex nature of the links between physical activity/sedentary behaviour and schizophrenia, and they suggest that increasing levels of physical activity might not be an effective intervention strategy for schizophrenia. Further research is needed to understand the impact of different types of physical activities on schizophrenia.

We also found weak evidence for the plausible protective effect of physical activity on bipolar disorder and PTSD, although earlier findings from observational and intervention studies have highlighted the potential beneficial impact of physical activity on these disorders.^7^ In addition, both PTSD and bipolar disorder were causally related to increased levels of physical activity. Increased physical activity might, thus, reflect psychopathological symptoms, such as high energy levels and disorganisation in mania or engagement in demanding activities to avoid re-experiencing in PTSD.

### Causal pathways between physical activity and eating disorders

People with eating disorders often engage in excessively high levels of physical activity in order to maximise energy expenditure and weight loss.^44^ Our results show that anorexia is causally related to higher levels of self-reported but not objectively assessed physical activity, suggesting that this disorder may have a greater impact on the subjective experience of physical activity than on the actual levels of physical activity undertaken. Another novel finding is that the odds of anorexia decreased by 30% for every 1-SD increase in the levels of sedentary behaviour. Therefore, enhancing sedentary behaviours could be an effective strategy to reduce the risk of anorexia.

### Causal pathways between physical activity and neurodevelopmental disorders

Initial evidence suggests that interventions involving physical activity might help to ameliorate certain symptoms of neurodevelopmental disorders such as ADHD and autism.^45–47^ Despite these findings, our results provide weak evidence for the plausible protective effect of physical activity on ADHD and autism. In the opposite direction of causation, we observed a causal effect of ADHD on higher levels of self-reported physical activity, while autism was causally linked to reduced levels of objectively assessed walking. Of note, the GWASs of ADHD and autism were largely conducted in children and young people, whereas the GWAS of physical activity was based on a sample of adults. These findings could therefore be inconclusive if the genetic determinants of physical activity in childhood are different from those in adulthood.

### Strengths and limitations

Our study has several strengths, including (i) the application of a genetically informed approach to strengthen causal inferences; (ii) the use of summary statistics drawn from the largest available GWASs with non-overlapping samples for each exposure-outcome relationship; (iii) the inclusion of different types of physical activity phenotypes based on both self-reported and objectively assessed data; (iv) a comprehensive assessment of the role of physical activity across a variety of mental health and substance use disorders; (v) the use of several sensitivity analyses and robust MR methods to ascertain the validity of key MR assumptions and assess the accuracy of the results; (vi) and the inclusion of a negative control outcome that, as expected, was not causally affected by any physical activity exposure included in our analysis.

Despite these strengths, the results should be interpreted in light of some limitations.^48^ First, it should be noted that causal effects from MR do not provide information on temporal patterns and should be interpreted as the lifetime effects of the liability to a particular risk factor. In addition, our measures of mental health/substance use disorders represent prevalent cases, so our results cannot clearly disentangle the role of physical activity in the prevention versus treatment of mental illness. Second, some of the included GWASs only identified few genome-wide significant SNPs associated with the exposures of interest (e.g., objectively assessed physical activity; see eTable1, Appendix 1), which could affect the power of the instruments. To address this, we used a second set of genetic instruments including top SNPs meeting a more relaxed p-value threshold, and we applied MR-RAPS to account for weak instrument bias. In addition, although we used the largest available GWASs, some were based on relatively small samples. Hence, the weak causal associations between physical activity and certain outcomes (e.g., autism, alcohol dependence) observed in our study could be explained by methodological issues related to the power of the instruments and the GWAS datasets, which might have increased the risk of false negative results (i.e., Type 2 error). Moreover, common SNPs usually explain a limited proportion of the total variance in complex traits, and their exact biological action is unclear to date. As such, we cannot rule out the possibility that pleiotropic mechanisms might have affected the main study results. Third, we found some evidence of a bidirectional causal relationship between sedentary behaviour and schizophrenia. However, a causal effect in both directions could be a product of violations of the second and third IV assumptions (see Figure1) (e.g., the genetics of personality or intelligence may influence both physical activity and mental illness) rather than indicating a true bidirectional relationship.^25^ Lastly, the genetic instruments for physical activity were all identified in the UK Biobank Study, which only includes adults aged 40 to 70 years and is not representative of the wider UK population. Furthermore, we do not have detailed information on the demographic characteristics of the participants included in the GWASs of mental health and substance use disorders. Therefore, our findings might not be generalisable to other populations and might have been affected by participation bias, which could influence both the strength and direction of the causal links between physical activity/sedentary behaviour and mental health/substance use disorders.

### Conclusions

In summary, this study capitalises on a genetically informed approach to test the plausible causal protective effects of physical activity on mental health and substance use disorders. Our findings suggest that physical activity has a protective effect on depression and cigarette smoking, whereas sedentary behaviour might reduce the risk of anorexia. Furthermore, they outline the likely impact of mental illness on physical activity levels, and they also point to the importance of considering different assessment methods, types, and intensity levels of physical activity in mental health research. Programmes to enhance physical activity may be an effective strategy to reduce the risk of depression and cigarette smoking. In contrast, the promotion of sedentary or light physical activities could help to reduce the risk of anorexia nervosa.

## Data Availability

The references for the data used in the study are provided in the manuscript.

## Acknowledgements

Summary-level data for the exposures and outcomes were drawn from large-scale GWASs or genetic consortia, including the UK Biobank, the Psychiatric Genomics Consortium, the GWAS and Sequencing Consortium of Alcohol and Nicotine Use, and the Early Growth Consortium.

## Code availability

The code of the statistical analyses can be accessed on GitHub: https://github.com/Ellie25moon/2-Sample-MR-study-of-physical-activity-and-mental-health-

## APPENDIX 1

### eMETHODS

**eTable1.**
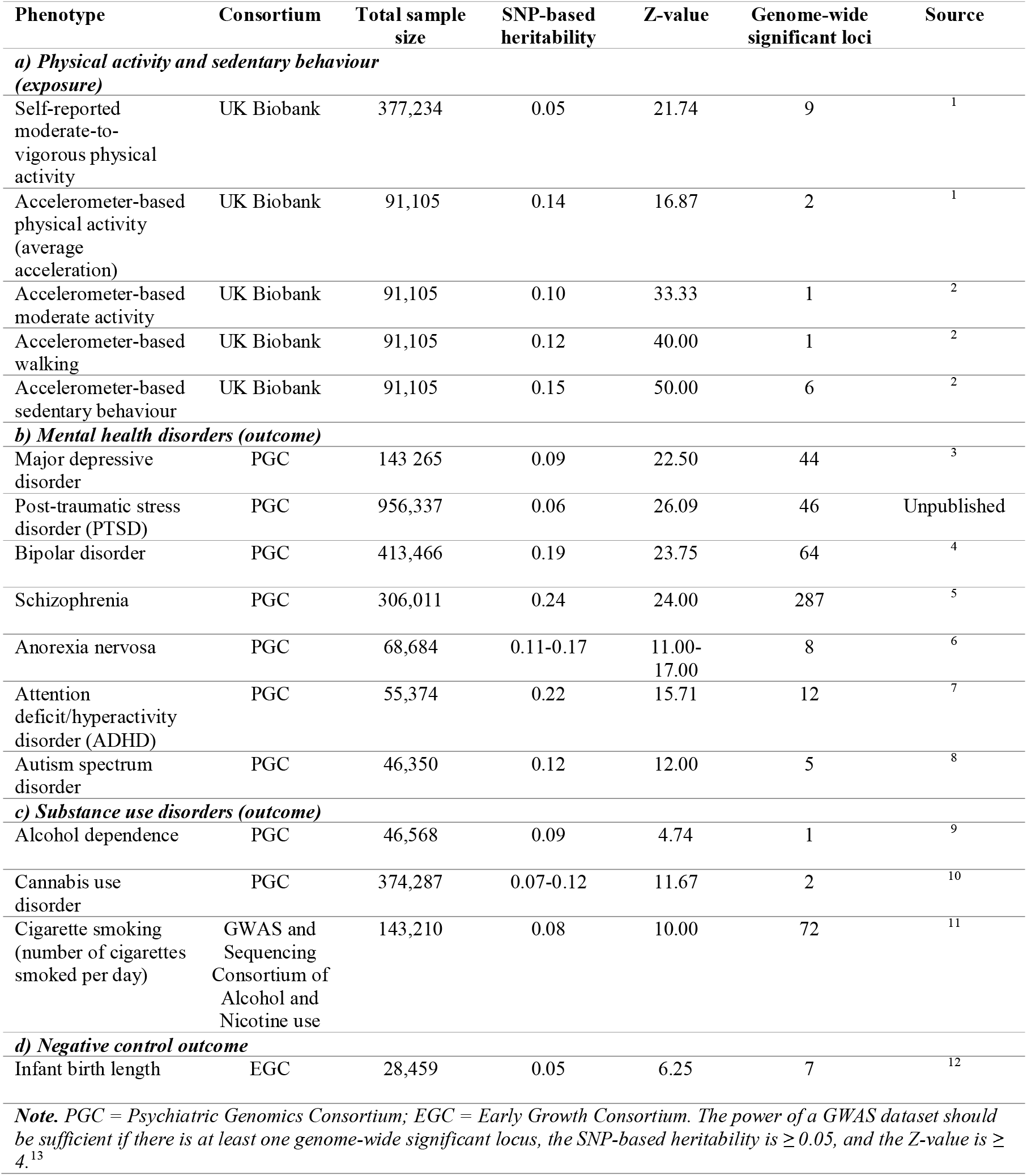
Overview of the GWAS datasets.

| Phenotype | Consortium | Total sample size | SNP-based heritability | Z-value | Genome-wide significant loci | Source |
| --- | --- | --- | --- | --- | --- | --- |
| **a) Physical activity and sedentary behaviour (exposure)** |  |  |  |  |  |  |
| Self-reported moderate-to-vigorous physical activity | UK Biobank | 377,234 | 0.05 | 21.74 | 9 | <sup>1</sup> |
| Accelerometer-based physical activity (average acceleration) | UK Biobank | 91,105 | 0.14 | 16.87 | 2 | <sup>1</sup> |
| Accelerometer-based moderate activity | UK Biobank | 91,105 | 0.10 | 33.33 | 1 | <sup>2</sup> |
| Accelerometer-based walking | UK Biobank | 91,105 | 0.12 | 40.00 | 1 | <sup>2</sup> |
| Accelerometer-based sedentary behaviour | UK Biobank | 91,105 | 0.15 | 50.00 | 6 | <sup>2</sup> |
| **b) Mental health disorders (outcome)** |  |  |  |  |  |  |
| Major depressive disorder | PGC | 143,265 | 0.09 | 22.50 | 44 | <sup>3</sup> |
| Post-traumatic stress disorder (PTSD) | PGC | 956,337 | 0.06 | 26.09 | 46 | Unpublished |
| Bipolar disorder | PGC | 413,466 | 0.19 | 23.75 | 64 | <sup>4</sup> |
| Schizophrenia | PGC | 306,011 | 0.24 | 24.00 | 287 | <sup>5</sup> |
| Anorexia nervosa | PGC | 68,684 | 0.11-0.17 | 11.00-17.00 | 8 | <sup>6</sup> |
| Attention deficit/hyperactivity disorder (ADHD) | PGC | 55,374 | 0.22 | 15.71 | 12 | <sup>7</sup> |
| Autism spectrum disorder | PGC | 46,350 | 0.12 | 12.00 | 5 | <sup>8</sup> |
| **c) Substance use disorders (outcome)** |  |  |  |  |  |  |
| Alcohol dependence | PGC | 46,568 | 0.09 | 4.74 | 1 | <sup>9</sup> |
| Cannabis use disorder | PGC | 374,287 | 0.07-0.12 | 11.67 | 2 | <sup>10</sup> |
| Cigarette smoking (number of cigarettes smoked per day) | GWAS and Sequencing Consortium of Alcohol and Nicotine use | 143,210 | 0.08 | 10.00 | 72 | <sup>11</sup> |
| **d) Negative control outcome** |  |  |  |  |  |  |
| Infant birth length | EGC | 28,459 | 0.05 | 6.25 | 7 | <sup>12</sup> |
| **Note.** PGC = Psychiatric Genomics Consortium; EGC = Early Growth Consortium. The power of a GWAS dataset should be sufficient if there is at least one genome-wide significant locus, the SNP-based heritability is $\geq 0.05$, and the Z-value is $\geq 4$.<sup>13</sup> | | | | | | |

#### Deviations from the pre-registered study protocol

Our original pre-registered study protocol also included anxiety disorders, obsessive compulsive disorder (OCD), and opioid dependence as primary outcomes/exposures. However, we have decided to exclude these disorders as their GWAS datasets may have limited power, as shown below.

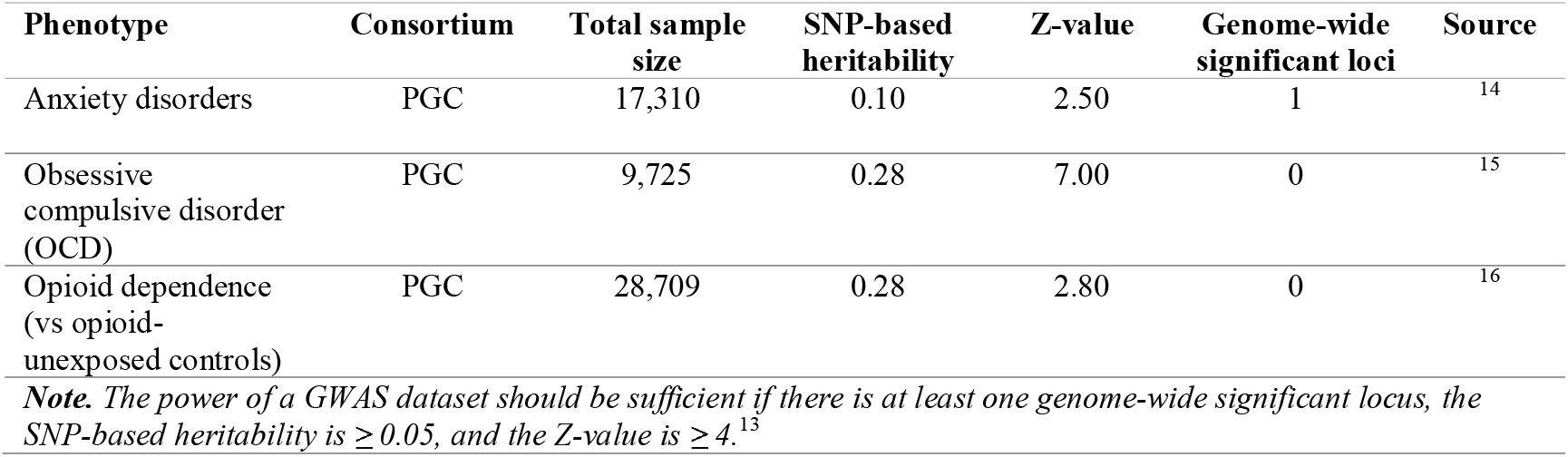

**eFigure1.**
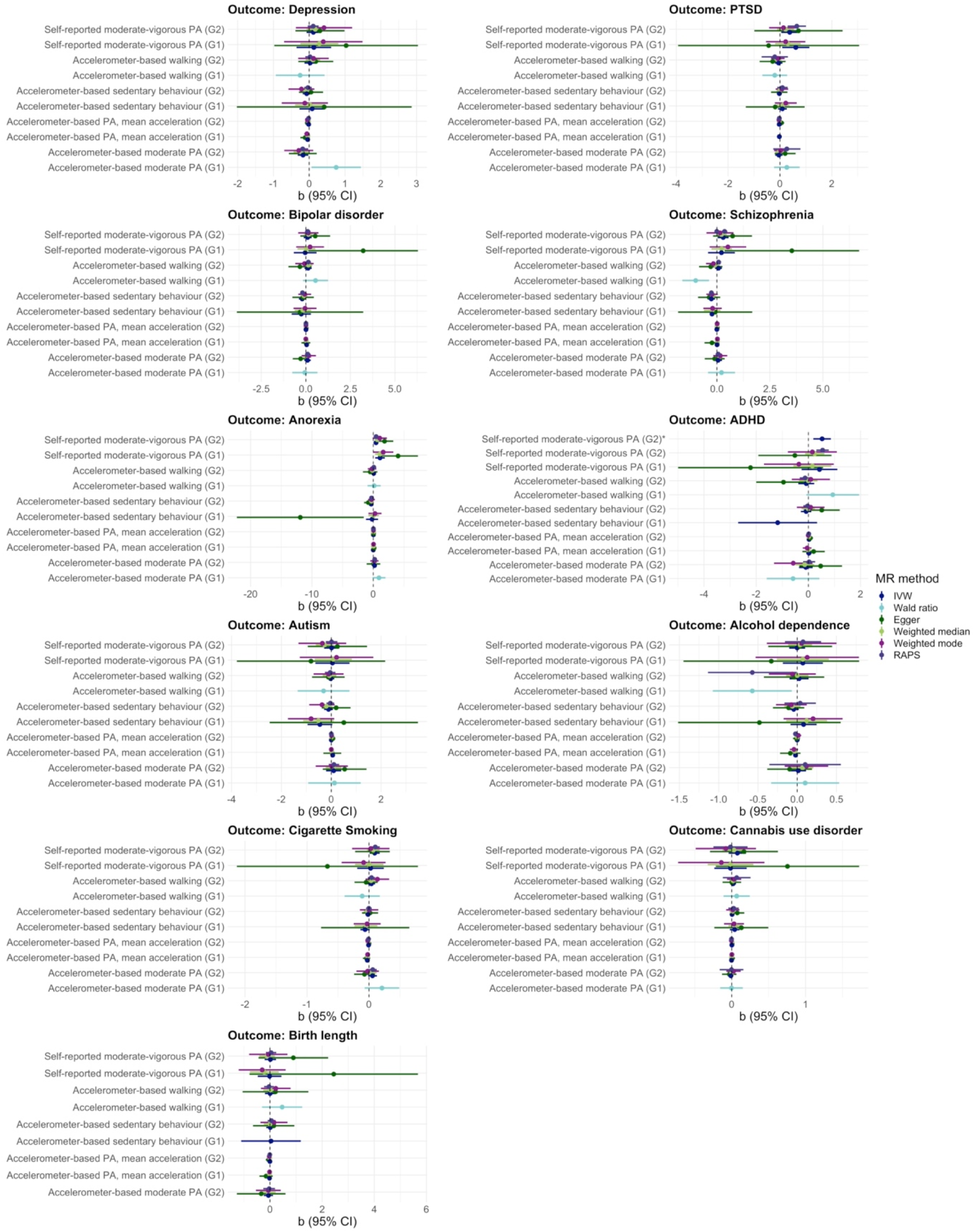
MR estimates and 95% confidence intervals for the causal relationships of physical activity and sedentary behaviour with mental health and substance use disorders. **Note**. MR = Mendelian randomisation; IVW = inverse variance weighted; RAPS = Robust adjusted profile score; CI = confidence interval; G1 = genome-wide significant genetic instrument (P < 5×10^−8^); G2 = more relaxed genetic instrument (P < 1×10^−6^); PA = physical activity.

**eFigure2.**
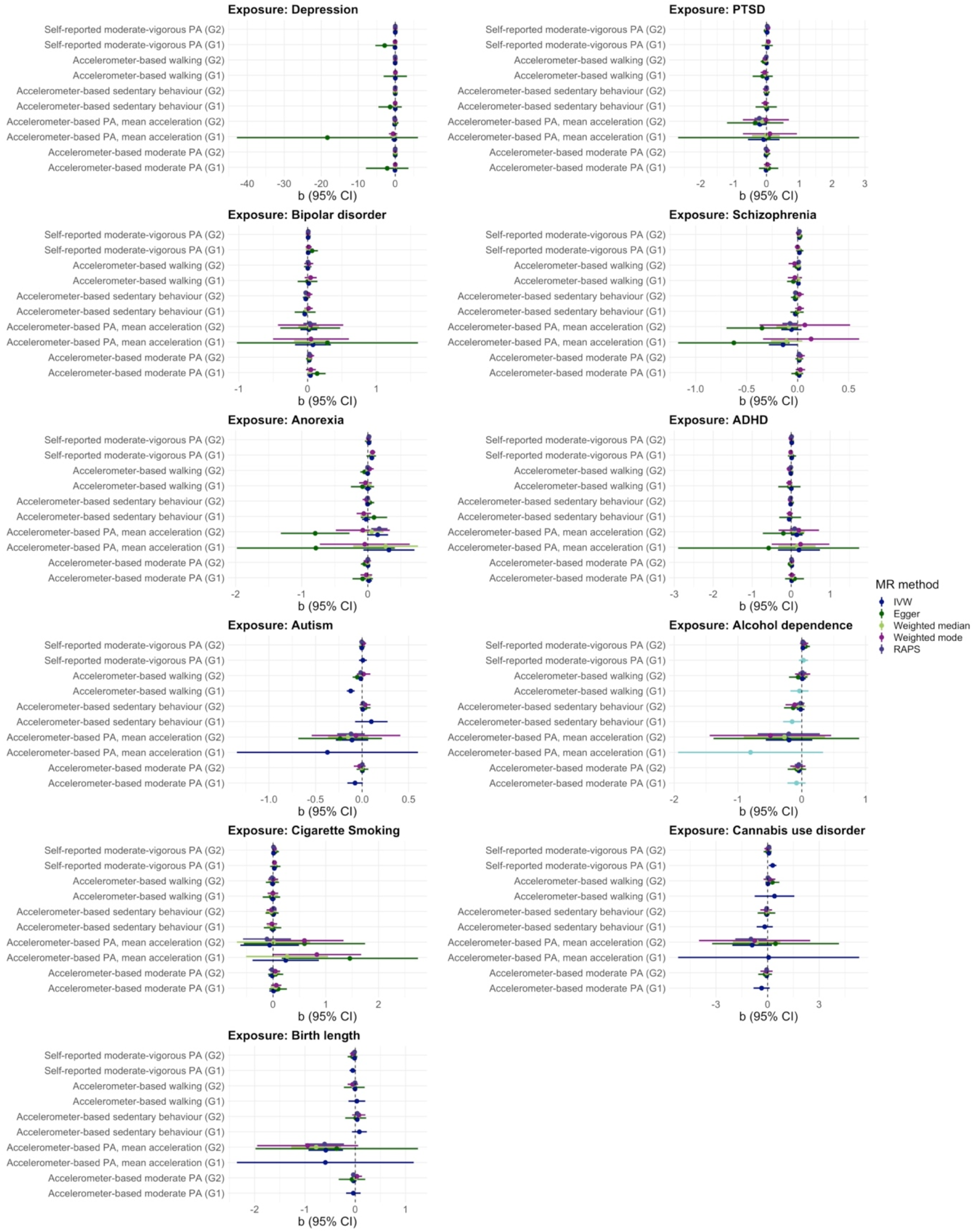
MR estimates and 95% confidence intervals for the causal relationships of mental health and substance use disorders with physical activity and sedentary behaviour. **Note**. MR = Mendelian randomisation; IVW = inverse variance weighted; RAPS = Robust adjusted profile score; CI = confidence interval; G1 = genome-wide significant genetic instrument (P < 5×10^−8^); G2 = more relaxed genetic instrument (P < 1×10^−6^); PA = physical activity.

